# The Emergence of SARS-CoV-2 Variant Lambda (C.37) in South America

**DOI:** 10.1101/2021.06.26.21259487

**Authors:** Pedro E. Romero, Alejandra Dávila-Barclay, Guillermo Salvatierra, Luis González, Diego Cuicapuza, Luis Solis, Pool Marcos-Carbajal, Janet Huancachoque, Lenin Maturrano, Pablo Tsukayama

## Abstract

We report the emergence of a novel lineage of SARS-CoV-2 in South America, termed C.37. It presents seven nonsynonymous mutations in the Spike gene (Δ247-253, G75V, T76I, L452Q, F490S, T859N) and a deletion in the ORF1a gene (Δ3675-3677) also found in variants of concern (VOCs) Alpha, Beta, and Gamma. Initially reported in Lima, Peru, in late December 2020, it now accounts for 97% of Peruvian public genomes in April 2021. It is expanding in Chile and Argentina, and there is evidence of onward transmission in Colombia, Ecuador, Mexico, the USA, Germany, and Israel. On June 15, 2021, the World Health Organization designated C.37 as Variant of Interest (VOI) Lambda.

## Announcement

The evolution of SARS-CoV-2 variants with potentially increased transmissibility, virulence, and resistance to antibody neutralization poses new challenges for the control of COVID-19 (1), particularly in low and middle-income countries (LMICs) where transmission remains high and vaccination progress is still incipient.

Peru has been severely hit by the COVID-19 pandemic: As of May 31, 2021, it had the highest rate of COVID-19 deaths globally relative to its population (180764 out of 33.38 million: ∼0.54% of the country’s population) (2). By June 2021, 1424 genome sequences from Peru were available on GISAID, comprising 64 circulating PANGO lineages (3). Routine genomic surveillance in early 2021 revealed a deep-branching sublineage of B.1.1.1, now classified as C.37 (Figure 1). It was first reported in Lima in December 2020 (1 of 192 genomes, 0.5%), expanding to 20.5%, 36.4%, 79.2%, and 96.6% in January, February, March, and April 2021, respectively (Figure 2). In contrast, Variants of Concern were detected less frequently over these four months in Peru: Alpha, n=7, 0.5%; Gamma, n=17, 1.2% (Figure 1).

**Figure 1.**
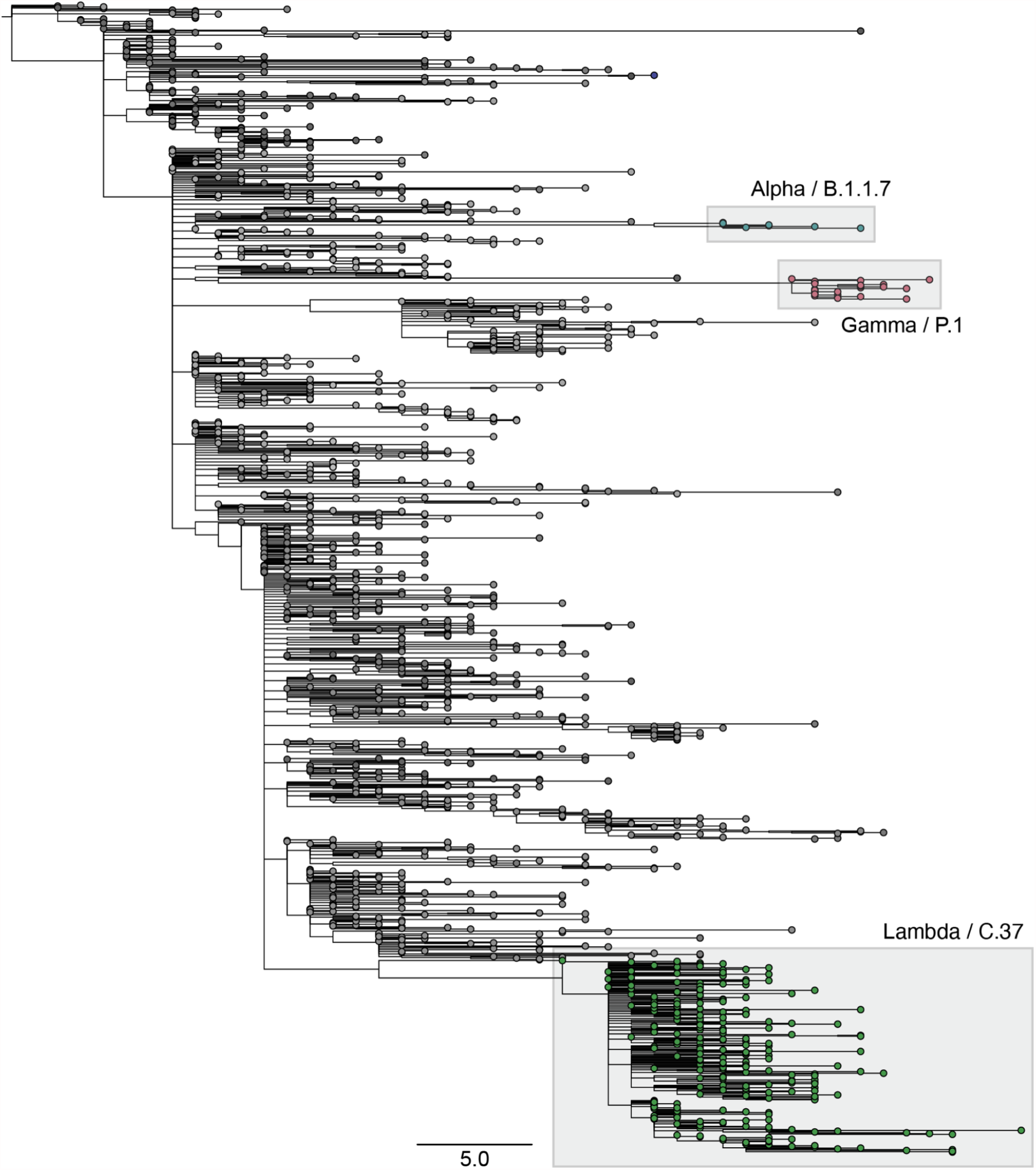
Maximum likelihood tree from 1424 SARS-CoV-2 genomes reported in Peru as of June 2021, highlighting variants Alpha (n=5), Gamma (n=26) and Lambda (n=239).

**Figure 2.**
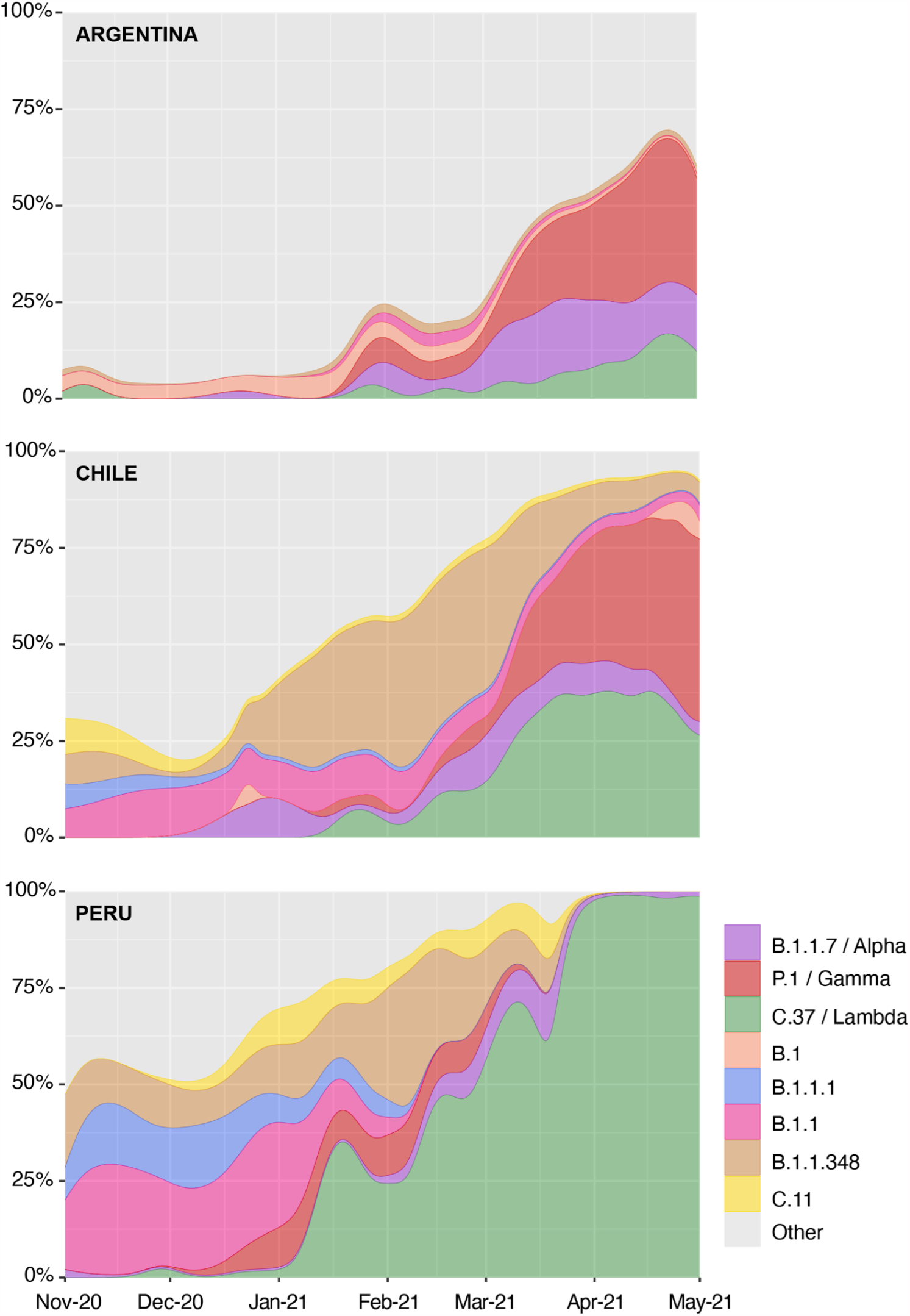
Relative frequencies of predominant SARS-CoV-2 lineages in Argentina, Chile, and Peru from November 2020 to May 2021.

C.37 contains a novel deletion (S:Δ247-253, located at the N-terminal domain) and six nonsynonymous mutations in the Spike gene (G75V, T76I, D614G, L452Q, F490S, T859N) (Table 1). Mutations L452Q and F490S both map to the Spike protein’s receptor-binding domain (RBD). While L452Q is almost exclusive to C.37, L452R is present in VOC Delta (B.1.617.2) and Variants of Interest (VOI) Epsilon (B.1.427/B.1.429) and Kappa (B.1.617.1) and is associated with increased affinity for the ACE2 receptor (4). F490S has been associated with reduced in vitro susceptibility to antibody neutralization (5,6). C.37 also displays the ORF1a:Δ3675–3677 deletion, found in VOCs Alpha, Beta, and Gamma (7).

**Table 1.** Nonsynonymous mutations present in SARS-CoV-2 lineage C.37 / Lambda.

| Gene | Amino acid |
| --- | --- |
| N | P13L, R203K, G204R, G214C, T366I |
| ORF1a | T1246I, P2287S, F2387V, L3201P, T3255I, G3278S, S3675-, G3676-, F3677- |
| ORF1b | P314L |
| ORF9b | P10S |
| S | G75V, T76I, R246N, S247-, Y248-, L249-, T250-, P251-, G252-, D253-, L452Q, F490S, D614G, T859N |

The earliest record of C.37 on GISAID is from Argentina in November 2020. By June 19, 2021, there were 1771 C.37 sequences from 25 countries, including Chile (n=670), USA (n=510), Peru (n=222), Argentina (n=86), Germany (n=79), Mexico (n=55), Spain (n=40), and Ecuador (n=39). Beyond Peru, C.37 has expanded rapidly in Chile and Argentina, reaching 33% and 12% of all sequenced genomes on GISAID by April 2021, respectively (Figure 2). The emergence of this lineage in Peru and its export to other countries is a current hypothesis, given its earlier detection and rise to nearly 100% of public sequences by April. We are sequencing additional Peruvian samples from October to December 2020 to confirm and date the origin of C.37.

Expansion of C.37 has occurred in South America in the presence of hundreds of circulating lineages and VOCs Alpha and Gamma (Figure 1B), suggesting increased transmissibility of this lineage. However, additional epidemiological data and analyses are needed to assess its transmission, virulence, and immune escape properties.

On June 15, 2021, the World Health Organization designated C.37 as VOI Lambda (8).

## Methods

Peruvian genomes were generated at Universidad Peruana Cayetano Heredia and Instituto Nacional de Salud using the Illumina COVIDseq or the Qiagen SARS-Cov-2 panel and QIAseq FX DNA library kits. Libraries were sequenced on Illumina MiSeq and NextSeq 550 instruments. Reads were processed and assembled with the Illumina DRAGEN COVID pipeline or a custom pipeline based on MEGAHIT v1.2.9. Assemblies with an average base coverage higher than 1000x were submitted to GISAID.

We downloaded SARS-CoV-2 genome assemblies from Argentina (n=2229), Chile (n=2564), and Peru (n=1424), available in GISAID by June 2021. Sequences were processed using the Nextstrain *augur* pipeline (9). Genomes were aligned against the Wuhan reference genome (NC_045512.2) using MAFFT v7.48. We then built a maximum likelihood phylogeny using IQTREE v.2.1.4. The tree was calibrated under a general time-reversible (GTR) model of nucleotide substitution, assuming a clock rate of 8×10^−4^. Two genomes from China (EPI_ISL_402123, EPI_ISL_406798) were used as the outgroup.

## Data Availability

All analyzed sequences were publicly available in GISAID at the time of submission. Raw Illumina reads from 350 Peruvian genomes sequenced at UPCH are available at NCBI BioProject PRJNA667090.

https://www.ncbi.nlm.nih.gov/bioproject/667090

## Data availability

All analyzed sequences were publicly available in GISAID at the time of manuscript submission. Raw Illumina reads from 350 Peruvian genomes sequenced at UPCH are available at NCBI BioProject PRJNA667090.

## Acknowledgments

The Institutional Review Board of Universidad Peruana Cayetano Heredia approved the project in June 2020 (E051-12-20).. We are funded by Fondo Nacional de Ciencia y Tecnología (FONDECYT) grants #046-2020, #022-2021, and Universidad Nacional Mayor de San Marcos grant #A2008007M. PER is supported by FONDECYT grant #034-2019. We thank our collaborators at Instituto Nacional de Salud for providing clinical specimens for sequencing. We acknowledge colleagues in the laboratories that generated and shared genetic sequence data via the GISAID Initiative, on which this study is based.

